# Use of the novel PanLeucogated CD4 test has saved over 600 million USD for South Africa’s HIV treatment programme: A 20-year retrospective costing analysis (2004 to 2024)

**DOI:** 10.64898/2026.02.18.26346526

**Authors:** Naseem Cassim, Wendy Susan Stevens, Deborah Kim Glencross, Lindi-Marie Coetzee

## Abstract

**Background:** In 2004, South Africa’s public health system faced the dual challenge of rapidly scaling up antiretroviral therapy (ART) while reducing the cost of laboratory monitoring. At the time, conventional CD4 testing methods were expensive, labour-intensive, and impractical for sustaining a national testing network. This study aimed to assess the financial impact and cost savings associated with the implementation of the PanLeucogated CD4 (PLG/CD4) enumeration method between 2004 and 2024 in the public-sector in South Africa.

**Methods:** A longitudinal cost analysis was conducted using annual test volumes and state tariffs for PLG/CD4 testing and the 4-colour CD3/CD4/CD8/CD45 T-cell enumeration reference method. Annual cost savings were calculated in United States Dollars (USD) by applying historical South African Rands (ZAR) to United States Dollars (USD) exchange rates. The state prices for tariff codes PLG/CD4 and the reference method were provided by calendar year in ZAR and converted to USD based on the prevailing exchange rate. The USD test prices were multiplied by annual test volumes. Cost savings were calculated by multiplying annual test volumes and the difference in test prices in USD (difference between PLG/CD4 and the reference method).

**Results:** There were 50,745,848 PLG/CD4 tests performed over 20-years. The cost-per-test of PLG/CD4 was consistently lower than the reference method, ranging from $4,06 to $9,40, compared to $13,06 to $28,21. Cumulative national savings amounted to USD 626 million. The peak annual savings of $64,6 million occurred in 2011, coinciding with the height of ART enrolment. Cost savings persisted despite a doubling in the exchange rate over the study period.

**Conclusion:** The PLG/CD4 implementation enabled cost-efficient, scalable, quality-assured CD4 testing as part of the national HIV response, reducing reliance on complex/costly technologies while improving coverage. These findings support the critical role of context-specific diagnostic innovation to strengthen health system resilience.

## Introduction

During the late 1990’s and early 2000’s, the political landscape required fortitude for people living with HIV (PLHIV).^[1-3]^ President Thabo Mbeki and Health Minister Manto Tshabalala-Msimang questioned the scientific consensus that HIV causes AIDS,^[4]^ leading to a delayed and inadequate response to the epidemic and reluctancy to provide antiretroviral therapy (ART)^[2]^, despite the availability of effective treatment.^[2]^ The very high costs of providing both ART and associated diagnostic laboratory monitoring, as well as the infrastructure and logistical challenges, further complicated and delayed the implementation of a national treatment programme.^[5]^ Advocacy groups such as Médecins Sans Frontières (MSF)^[6]^ and the Treatment Action Campaign (TAC)^[1, 7]^ actively campaigned against the AIDS denialist-stance, while raising awareness and fighting for the availability of affordability ART as a vital component of access to comprehensive HIV care.

In 2004, it was estimated that over 5 million South Africans of a total population of 46 million were living with HIV, revealing a national prevalence rate of 11%.^[3]^ Modelling work^[8]^ undertaken in 2006, suggested that the epidemic was set to overwhelm local health care services; the provision of ART, on the other hand, would substantially reduce AIDS deaths and curtail predicted health care costs.^[8]^ By 2010, the predicted likelihood of AIDS deaths were roughly 380,000 per annum, compared to an estimated 495,000 without the provision of ART.^[3]^

By 2002, the HIV pandemic was rapidly emerging.^[9-12]^ with calls to lower the costs of a treatment programme and secure better prices for ART.^[2]^ Advocacy for extended, widescale and affordable laboratory services also began,^[13-15]^ including ‘safety’ tests like full blood counts and liver function tests, amongst others.^[16]^ The CD4 cell counts, that provide a measure of disease progression in HIV and the immediate risk of death,^[17-20]^ were needed to identify PLHIV eligible for care, alongside HIV viral load (VL) testing needed to assess viral suppression. At the time, multiple studies analysed the costs of extending ART in the public sector using different programmatic options.^[21-23]^ Geffen *et al* reported that the cost of VL and CD4 monitoring in 2003 would be R707,40 South African Rand (ZAR), over and above the monthly cost of R355 for ART.^[22]^ This additional burden on the overall costs of a national treatment programme were substantial, considering that the Operational Plan for Comprehensive HIV and AIDS Care, Management and Treatment for South Africa aimed to initiate 1,470,510 PLHIV on ART between 2003/4 and 2008/9. In 2008/9, the budget for the operational plan was 4,474 million ZAR, of which 806 million (18%) was allocated for laboratory testing.^[23, 24]^ CD4 and VL testing, therefore, appeared unaffordable for widescale adoption in a national treatment programme.^[24]^

In addition to the high costs of diagnostic monitoring, expertise was urgently needed across all levels of services.^[14]^ In the National Health Laboratory Service (NHLS), at this time, technical staff were largely unprepared for the anticipated high volumes of PLHIV that were expected to access care.^[14]^ Laboratories were also considered ill-equipped to adequately handle the massive, anticipated scale-up of diagnostic services; the lack of appropriate technical skills to perform these new technologies, across all levels of service, also posed a challenge.^[13, 16]^. Technical expertise for specialised pathology testing at the time, especially for CD4 counts or VL assays, were also restricted to a few academic institutions. To equitably scale-up services, the entire nationwide existing laboratory infrastructure needed upgrading and staff across the organisation needed to be trained.^[13]^

At the start of the HIV epidemic, CD4 enumeration was technically demanding and costly. Early Centers for Disease Control and Prevention (CDC) guidelines (1992, 1994) recommended flow cytometry to perform the ‘lymphosum’ or 6/2 method, a complex, multi-reagent diagnostic panel system, necessitated advanced flow cytometry expertise, extensive manual preparation and laborious processes to ensure quality control.^[25-29]^ The method was poorly suited to low-resource settings like South Africa, where flow cytometry infrastructure and trained personnel were largely absent in routine laboratory services.^[30, 31]^

Efforts to simplify and automate CD4 testing led to the introduction of several streamlined “gating” methods designed to reduce reagent use and technical complexity.^[32-35]^ However, these approaches faced criticism for inconsistent quality control, and failed to achieve broad adoption in international guidelines. With the launch of South Africa’s public ART programme in 2004, there was an urgent need to establish a scalable, cost-effective national CD4 testing service.^[9, 10, 14]^

The novel PanLeucogated CD4 (PLG/CD4) method, a local South African innovation, was used to replace complex lymphocyte-based enumeration with a simplified two-reagent assay (CD4 and CD45) and save costs.^[36, 37]^ The simplified method was a paradigm shift in the way CD4 enumeration had been previously undertaken, enabling scale-up across the NHLS into routine laboratories across the country. PLG/CD4 was adopted nationally in 2004, after having being patented locally in 2001 and internationally by 2011 by the NHLS.^[16, 38]^ Details about the history of CD4 counting and PLG/CD4 are published elsewhere.^[16, 33, 38-40]^

The objective of this study was to document the financial impact and cost savings of PLG/CD4 in South Africa since the onset of the HIV programme between 2004 and 2024 by assessing both annual and total cost savings.

## Materials and Methods

### Ethical considerations

Ethical clearance for this study was obtained from the University of the Witwatersrand (M220163). Only test volumes and costs were used; our study did not require the use of any patient identifiers. Test volumes and state price data were originally extracted between 10/05/2024 and 31/12/2024. The retrospective extract included historical information from 01/01/2004 to 31/12/2024.

### Context

PLG/CD4 testing during the study period was offered using the following Beckman Coulter (Brea, CA, USA) flow cytometers: (i) XL-MCL (2004-2014), (ii) FC500 MPL/CellMek (2014-2023) and (iii) Aquios CL cytometers (2017 onward).^[41-43]^ Equipment was provided in a placement agreement using the national tender processes, in line with national supply chain procedures^[44]^ and included all maintenance and service. Irrespective of the flow cytometry instrument platform provided by Beckman Coulter, all laboratories utilised the standardised PLG/CD4 reagents and national standard operating procedures. The Beckman Coulter CYTOSTAT Tetrachrome 4-colour reagent, as an alternative best cost-saving option at the time, was used for comparison, as it was commercially available in South Africa since 2002.

### Study design

The cross-sectional study design was used to analyse CD4 test volumes and costs by calendar years 2004 to 2024.

### Cost-saving analysis

The study conducted a comprehensive micro-costing analysis of all inputs associated with PLG/CD4 and CD3/CD4/CD8/CD45 testing. The state prices for tariff codes 2485 (PLG/CD4) and 2486 (CYTOSTAT Tetrachrome) were provided by the NHLS Finance Department by calendar year.^[45]^ The state price in ZAR was converted to United States Dollars (USD) using the prevailing annual average exchange rates sourced from Nedbank (Johannesburg, South Africa).^[46]^ The state prices accounted for the following cost categories (equipment (procured using a placement agreement and included in reagent costs), reagents, test consumables, specimen transportation, waste disposal and overhead costs. The state price is the total charge for services offered to the public health sector.^[30]^ No outright purchase of flow cytometers were required for CD4 testing. The annual cost saving was calculated as the difference between the cost of PLG/CD4 and the alternative closest cost-saving method of CD3/CD4/CD8/CD45 (Beckman Coulter CYTOSTAT Tetrachrome). The cost saving per test was calculated per year (difference between tariff codes 2486 and 2485). The cost per test is the actual charge for CD4 services offered by the NHLS, based on a fee for service funding model.^[30]^ The public health sector and foreign governments that purchase laboratory tests from NHLS must pay the prescribed fees as determined after consultation with the Minister of Health.^[30]^ The state price should be inclusive of reagents, labour, overhead and fixed costs.^[30]^ However, cross-subsidisation across all tests offered by the NHLS are a factor that should be considered. The NHLS collects fees from all tests performed nationally. Routine tests generate steady income, which subsidises advanced or costly assays.^[30, 47]^ However, this should not be a factor for priority testing. Annual test volumes were provided by the NHLS Corporate Data Warehouse (CDW). The total annual cost savings were calculated by multiplying test volumes and cost savings in USD. The total cost saving over the study period was also determined. The data was prepared and analysed using Microsoft Excel (Microsoft Corporation, Redmond, WA, USA). Full calculations of the costs of a 6/2 lymphosum panel comparison to PLG/CD4 savings were not done (as the 6/2 panel option could never be a consideration for practical or financial reasons and as such, no tariff code pricing was ever implemented).

## Results

The annual ZAR/USD average exchange rate ranged from 6,3806 in 2005 to 18,4519 by 2023.^[46]^ The cost-per-test for a PLG/CD4 test ranged from $4,06 (2016) to $9,40 (2005), compared to between $13,06 (2016) and $28,21 (2005) for the CYTOSTAT Tetrachrome method (Table 1). Decreases in USD/ZAR exchange rates from 2005 to 2016 were noted. The price in ZAR, as paid by the National Department of Health (NDOH) to the NHLS for a PLG/CD4 test, increased modestly over the study period. Between 2004 and 2008, the annual negotiations, of the NDOH with the NHLS, kept the price of the state CD4 service consistent at R60,00 for just under 5 years. Over the 20-year period, the state price of PLG/CD4 testing increased by R21,93. This is in keeping with a portfolio committee statement by the NHLS in 2005/06 that tariff rates for tests would be kept low by the NHLS in comparison to private pathologists.^[48]^ In ZAR, the cost of the PLG/CD4 test reported a range of R57,48 (2012) to R79,41 (2021), with an increase of 38,15% over the study period. For the CYTOSTAT Tetrachrome method, the cost-per-test ranged from R177,97 (2009) to R255,30 (2023) (Fig 1), by comparison reported a higher 43,45% increase. Data is reported for 50,745,848 PLG/CD4 tests, with annual volumes increasing from 45,081 in 2004, peaking at 3,933,642 by 2011 (Table 1). The annual cost savings ranged from $837,211 in 2004 to $64,640,892 by 2011.^[46]^ Between 2004 and 2024, the total cost savings was $626,778,870.

**Table 1:** Analysis of the annual cost savings of using PLG/CD4 between 2004 and 2024 compared to multi-colour (4-colour CD3/CD4/CD8/CD45 T-cell) T-cell enumeration. The analysis is based on the state price list from the National Health Laboratory Service (NHLS) and ZAR/USD annual average exchange rates.

| Year | Test | Cost per Test (ZAR) |  | Cost per Test (USD) |  | Exchange Rate | Saving (USD)<br>per test B | Total annual<br>savings (USD) A*B |
| --- | --- | --- | --- | --- | --- | --- | --- | --- |
|  | Volumes<br>n= (%) A | PLG/CD4 | Multi-colour T-cell<br>enumeration | PLG/CD4 | Multi-colour T-cell enumeration | Rate<sup>§</sup> |  |  |
| 2004 | 45,081 (0.1) | R60,00 | R180,00 | \$9,29 | \$27,86 | 6,4616 | \$18,57 | \$837,211 |
| 2005 | 78,079 (0.2) | R60,00 | R180,00 | \$9,40 | \$28,21 | 6,3806 | \$18,81 | \$1,468,432 |
| 2006 | 183,853 (0.4) | R60,00 | R180,00 | \$8,85 | \$26,55 | 6,7796 | \$17,70 | \$3,254,227 |
| 2007 | 663,490 (1.4) | R60,00 | R180,00 | \$8,50 | \$25,49 | 7,0625 | \$16,99 | \$11,273,458 |
| 2008 | 1,917,889 (3.9) | R60,00 | R180,00 | \$7,26 | \$21,78 | 8,2640 | \$14,52 | \$27,849,308 |
| 2009 | 2,309,818 (4.7) | R63,60 | R177,97 | \$7,53 | \$21,08 | 8,4440 | \$13,54 | \$31,285,396 |
| 2010 | 3,541,341 (7.3) | R60,28 | R179,75 | \$8,22 | \$24,51 | 7,3324 | \$16,29 | \$57,700,618 |
| 2011 | 3,933,642 (8.1) | R60,28 | R179,75 | \$8,29 | \$24,72 | 7,2702 | \$16,43 | \$64,640,892 |
| 2012 | 3,858,025 (7.9) | R57,56 | R185,04 | \$7,00 | \$22,51 | 8,2206 | \$15,51 | \$59,827,875 |
| 2013 | 3,679,464 (7.6) | R59,86 | R192,44 | \$6,20 | \$19,92 | 9,6588 | \$13,73 | \$50,505,584 |
| 2014 | 3,783,437 (7.8) | R59,86 | R192,44 | \$5,51 | \$17,72 | 10,8605 | \$12,21 | \$46,186,463 |
| 2015 | 3,573,447 (7.3) | R57,48 | R184,80 | \$4,50 | \$14,47 | 12,7721 | \$9,97 | \$35,622,276 |
| 2016 | 3,379,793 (6.9) | R59,78 | R192,19 | \$4,06 | \$13,06 | 14,7129 | \$9,00 | \$30,416,736 |
| 2017 | 3,057,313 (6.3) | R62,89 | R202,18 | \$4,72 | \$15,18 | 13,3209 | \$10,46 | \$31,968,795 |
| 2018 | 2,816,065 (5.8) | R66,47 | R213,70 | \$5,02 | \$16,13 | 13,2471 | \$11,11 | \$31,298,114 |
| 2019 | 2,804,525 (5.8) | R69,79 | R224,39 | \$4,83 | \$15,52 | 14,4566 | \$10,69 | \$29,991,808 |
| 2020 | 2,429,891 (5.0) | R72,51 | R233,14 | \$4,40 | \$14,16 | 16,4651 | \$9,76 | \$23,705,498 |
| 2021 | 2,261,463 (4.5) | R74,69 | R240,13 | \$5,05 | \$16,23 | 14,7955 | \$11,18 | \$25,287,178 |
| 2022 | 2,233,598 (4.4) | R77,85 | R250,29 | \$4,76 | \$15,29 | 16,3720 | \$10,53 | \$23,525,632 |
| 2023 | 2,135,648 (4.2) | R79,41 | R255,30 | \$4,30 | \$13,84 | 18,4519 | \$9,53 | \$20,357,748 |
| 2024 | 2,059,986 (4.1) | R79,41 | R255,30 | \$4,33 | \$13,93 | 18,3221 | \$9,60 | \$19,775,623 |
| **Total** | **50,745,848 (100.00)** | | | | | | | **\$626,778,870** |

<sup>§</sup>Exchange rates obtained from Nedbank<sup>[46]</sup> USD: United States Dollars USD: ZAR: South African Rands PLG: PanLeucogated NHLS: National Health Laboratory Service

**Fig 1:**
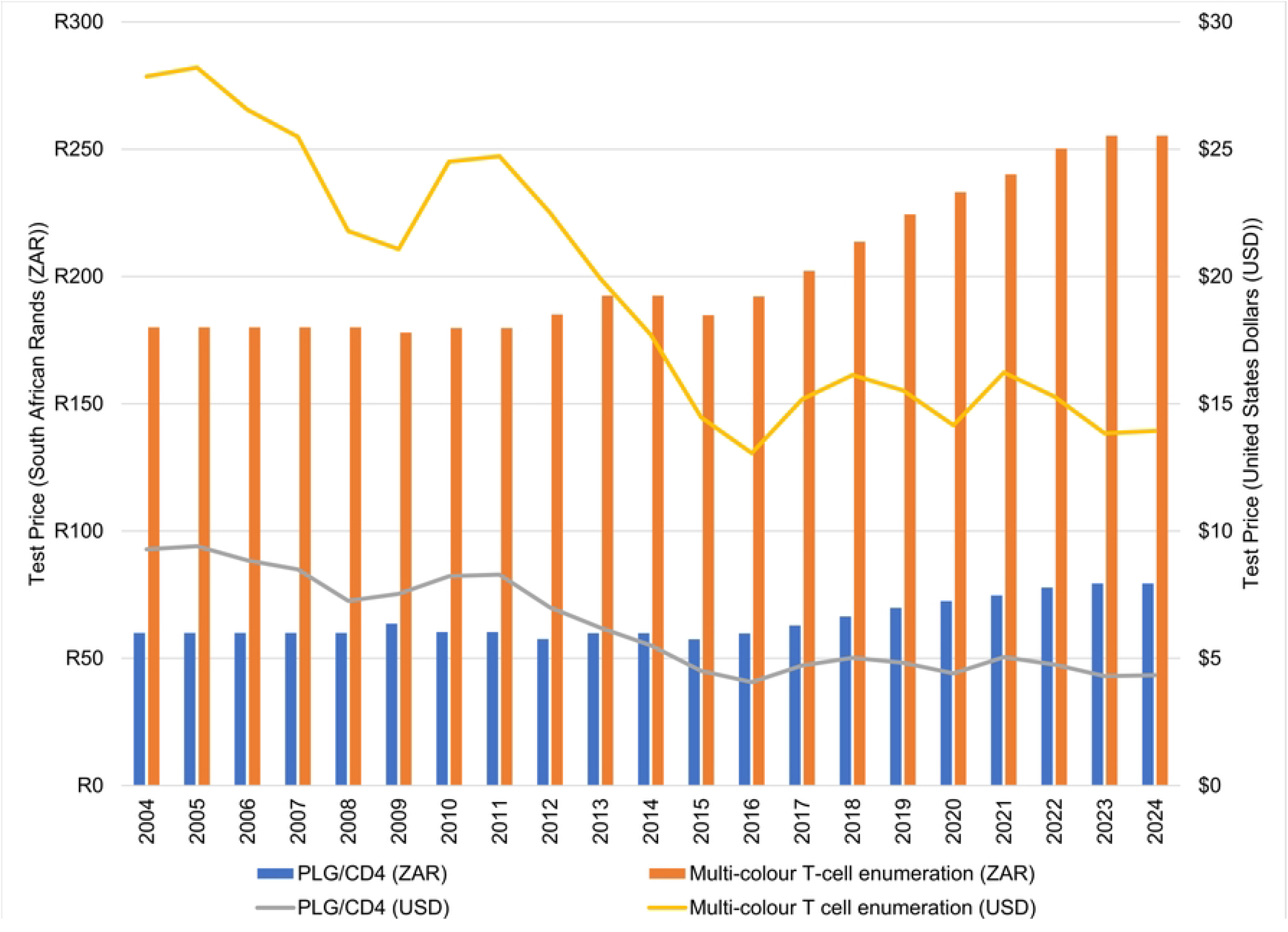
Test price for PLG/CD4 and multi-colour T-cell enumeration (4-colour CD3/CD4/CD8/CD45 T-cell) enumeration between 2004 and 2024 reported in both ZAR and USD. The analysis is based on the state prices provided by the National Health Laboratory Service (NHLS) and corresponding annual average ZAR/USD exchange rates. USD: United States Dollars ZAR: South African Rands PLG: PanLeucogated NHLS: National Health Laboratory Service

## Discussion

This study presents a comprehensive bottom-up micro-costing analysis of PLG/CD4 testing in South Africa’s public health sector. This testing was provided by the NHLS laboratory network, formed in 2000 as a government sector parastatal^[30]^ and incorporating all existing laboratories across nine South African provinces. This service was, and continues to be, the preferred provider of the NDOH for laboratory services offering a wide repertoire of both essential and specialist pathology testing to the public health clinical sector.^[49]^ Analysis of two decades of laboratory data presented here demonstrates that the implementation of the local PLG/CD4 test solution, developed in the face of significantly costly and complicated prevailing guideline-suggested test methods, has resulted in a total cost saving of over USD $626 million (7 billion ZAR) between 2004 and 2024. Annual cost savings peaked in 2011 at approximately $64.6 million, coinciding with a phase of high patient enrolment of the HIV programme when the highest number of CD4 tests were done in a single year; the total number of PLHIV starting ART that year was estimated to be 656,952 by the Thembisa model.^[50]^ Furthermore, this cost-saving was obtained during a period when the ZAR/USD exchange rate reported an unfavourable 2-fold change. These findings underscore the critical role that strategic diagnostic innovation has played in enabling the scale-up of comprehensive HIV services across South Africa. This simplified PLG/CD4 test not only enabled expanded access but further preserved diagnostic integrity whilst drastically lowering costs. The appropriateness, accessibility and affordability of PLG/CD4 testing thus played a pivotal role in the broader rollout of ART within a tiered laboratory service model.^[51, 52]^

While many studies have documented the clinical importance of CD4 monitoring in early HIV programmes, this study is the first to quantify the cumulative national cost savings of a specific CD4 enumeration methodology over such an extended period. A key strength of this study lies in its longitudinal scope and real-world data basis, using state tariff pricing and national test volumes from the NHLS CDW, an important national database of curated laboratory data for the entire public health sector in South Africa. ^[51-54]^ This work also demonstrates how consistent use of a dedicated methodology, successfully adapted onto new-generation platforms,^[36, 41, 52, 55, 56]^ and implemented across a national laboratory network for over 20 years, including implementation into smaller district and community laboratories with no previous exposure to flow cytometry,^[51, 52, 57, 58]^ can strengthen a national health programme.

Diagnostic innovation tailored to local constraints can drive substantial health system efficiencies. PLG/CD4 was not only a technological improvement, but a strategic enabler of national treatment access. The work shows that public sector laboratories are fully capable of innovation and can grow their ideas in partnership with academic and activist stakeholders, to solve infrastructural and cost-related health care challenges, and at scale. Further, as countries consider the future role of CD4 monitoring amid “test and treat” and universal ART guidelines, the value of low-cost, robust, accessible CD4 methods remains relevant, especially for identifying advanced HIV disease^[59-61]^, supporting differentiated service delivery models, and guiding clinical management where VL testing is limited.^[62-68]^

Further research is warranted to assess the full cost-effectiveness of the PLG/CD4 innovation. Modelling that includes clinical outcomes and health systems impact could complement this analysis. In addition, adapting the PLG/CD4 model to other low- and middle-income countries with similar infrastructural limitations could have broader global implications for diagnostic cost containment and health equity.

## Limitations

Several limitations merit discussion. First, the analysis focuses solely on direct diagnostic cost savings associated and does not include non-diagnostic benefits such as improved health outcomes due to timely ART initiation or reduced disease burden from delayed progression. The work also does not consider possible downstream savings linked to reduced laboratory staff time and reagent storage^[51]^, improved workflow^[55, 69, 70]^, improved turnaround time (TAT)^[71-73]^ or logistic improvements that contribute to service delivery^[74]^ that were developed to support the national CD4 programme. The economic impact of related programmes directly linked to, and supported by, the South African CD4 services for the detection of opportunistic infection is described elsewhere.^[75, 76]^ Specifically, in this regard, the PLG/CD4 programme has provided the laboratory network and platform to rollout the national reflexed Cryptococcal antigenaemia screening service.^[61, 75-78]^ Savings linked to these initiatives, and the economic impact of lives saved, were beyond the scope of this analysis.

Secondly, this study compares PLG/CD4 exclusively to four-colour CD3/CD4/CD8/CD45 method, considered the alternative prevailing cost-cutting alternative at the start of the South African HIV programme, and not to the ‘lymphosum/6/2 method.^[25]^ Although point of care (POC) enumeration technologies, used elsewhere in resource-poor settings and including PointcareNOW (Marlborough, MA, USA)^[79]^, Alere ^[80]^PIMA^[81]^ (Abbott, Chicago, IL, USA), Becton Dickinson (BD) FACSCount^[82]^ or BD Presto ^[83]^, amongst others, are an important component to extend services to hard-to-reach areas ^[52]^ as well as to provide the clinical benefit of same-day results.^[80]^ Studies have shown that POC testing substantially reduced attrition between CD4 testing and ART initiation when compared to conventional the laboratory-based standard of care.^[84, 85]^ POC CD4 allows for it allows timely, correct treatment and counselling in real time. This also a many additional costs for the patient and the healthcare system.^[86]^ This study did not conduct a comparative analysis of the programmatic costs of widescale implementation.

Thirdly, data is only reported for South Africa. The programme may have lower CD4 costs overall due to the national supply chain policy and the Public Finance Management Act ^[44, 87]^ that adopts a placement arrangement for testing platforms, and which includes service and maintenance costs based on economies of scale, due to the substantively higher volumes of tests performed across the country. In other regional settings, the cost savings may not be as significant where an outright purchase option of laboratory equipment is adopted (in other words, where flow cytometry instruments are bought outright) and test volumes are much lower.

Finally, cost was the primary metric; this study does not explicitly address the savings associated with improved diagnostic performance characteristics, e.g., accuracy, precision, or clinical impact of a more reliable test, as reported for PLG/CD4.^[16, 36, 56]^

## Conclusion

This study demonstrates that the adoption of the PLG/CD4 enumeration method, developed to meet specific needs for laboratory CD4 testing in a resource poor setting, and implemented locally in South Africa, resulted in substantial cost savings, exceeding USD $626 million (over 7 billion ZAR) for the country’s HIV programme between 2004 and 2024. These savings were achieved despite significant fluctuations in currency exchange rates while maintaining CD4 test accuracy, scalability, and improving test quality. Beyond the financial impact, PLG/CD4 enabled decentralised CD4 testing and supported rapid expansion of ART services across the country, including in underserved and remote regions.

The success of PLG/CD4 provides a compelling model for future investment in context-appropriate health technologies across the region. The approach exemplifies how locally driven diagnostic innovation, tailored to health system constraints and resource settings, can produce high-impact solutions with lasting national benefit. This is especially important given the evolving global funding priorities and increasing pressure on domestic health budgets, recently faced with the widescale withdrawal of grant-based funding.^[88]^ Political and financial regional commitment to research that provides opportunity for African scientists to develop solutions that address regional health issues will be essential for future support of national HIV, and other health programmes across the region, to ensure meeting the Joint United Nations Programme on HIV/AIDS (UNAIDS) 95-95-95 targets^[89]^ and ensure long-term regional programme resilience. Lastly, the authors hope that the story of implementation PLG/CD4 will inspire the next generation of young African scientists, researchers and clinicians to confront current and future health challenges with the same championing of cause and problem-solving approach that led to the proudly South African development of PLG/CD4.

## Acknowledgements

The authors would like to thank Niresh Booplal and Silence Ndlovu.

## Data availability statement

All cost data has been shared in this publication.

## Author Contributions

NC & LC (Developed and executed research, conducted data analysis, prepared the first draft and provided editorial input). LC, WSS & DKG (Editorial input and project leader)

## Disclaimer

The views expressed in this manuscript are those of the author(s) and not those of the University of the Witwatersrand or the NHLS.

## References

1. Grebe E. The Treatment Action Campaign’s Struggle for AIDS Treatment in South Africa: Coalition-building Through Networks. Journal of Southern African Studies. 2011;37(4):849–68. doi: 10.1080/03057070.2011.608271

2. Heywood M. South Africa’s Treatment Action Campaign: Combining Law and Social Mobilization to Realize the Right to Health. Journal of Human Rights Practice. 2009;1(1):14–36. doi: 10.1093/jhuman/hun006

3. Dorrington R, Bradshaw D, Johnson L, Budlender D. National Indicators for 2004: Centre for Actuarial Research, South African Medical Research Council, Actuarial Society of South Africa 2004 [Available from: https://www.hst.org.za/publications/NonHST%20Publications/HIVIndicators_2004.pdf]

4. Fassin D, Schneider H. The politics of AIDS in South Africa: beyond the controversies. BMJ. 2003;326(7387):495–7. doi: 10.1136/bmj.326.7387.495

5. Schoub BD, Smith A, Johnson S, Martin D, Lyons S, Padayachee G, et al. Considerations on the further expansion of the AIDS epidemic in South Africa--1990. S Afr Med J. 1990;77(12):613–8. doi:

6. Médecins Sans Frontières (MSF). 1999-2019: 20 Years of Advocacy in Action 2025 [Available from: https://msfaccess.org/1999-2019-20-years-advocacy-action]

7. Constitutional Court of South Africa. Minister of Health and Others v Treatment Action Campaign and Others (No 2) (CCT8/02) [2002] ZACC 15; 2002 (5) SA 721 (CC); 2002 (10) BCLR 1033 (CC) (5 July 2002). Constitutional Court of South Africa; 2002.

8. Johnson LF, Dorrington RE. Modelling the demographic impact of HIV/AIDS in South Africa and the likely impact of interventions. Demographic Research. 2006;14:541–74. doi: 10.4054/DemRes.2006.14.22

9. Broomberg J, Steinberg M, Masobe P, Behr G, editors. AIDS in South Africa: The Demographic and Economic Implications. University of the Witwatersrand: Centre for Health Policy; 199. Editors’ Summary of the Brookings Papers on Economic Activity – 2001 No 1; 2001; Washington, DC, USA: The Brookings Institution.

10. McIntyre J. HIV/AIDS in South Africa--a relentless progression? S Afr Med J. 1996;86(1):27–8. doi:

11. Sher R. HIV infection in South Africa, 1982-1988--a review. S Afr Med J. 1989;76(7):314–8. doi:

12. Schoub BD. The AIDS epidemic in South Africa--perceptions and realities. S Afr Med J. 1990;77(12):607–8. doi:

13. Glencross D, Mendelow B, Stevens W. Laboratory monitoring of HIV/AIDS in a resource-poor setting: AIDS forum: SAMJ forum. South African Medical Journal. 2003;93(4):262–3. doi:

14. Stevens W, Fiscus SA, Glencross D, Rehkviashvili N, Sherman G, Wallis C, et al. Affordable HIV diagnosis and monitoring for scaling up ARV treatment programmes. Southern African Journal of HIV Medicine. 2005;2005(20):38–41. doi: 10.10520/EJC65340

15. Treatment Action Campaign (TAC). Working for Access to Quality Healthcare in South Africa 1998 [Available from: https://tac.org.za/]

16. Glencross DK. Panleucogated (PLG) CD4 HIV Immune Monitoring: A difference by disruption: University of Witwatersrand; 2021.

17. Giorgi JV, Detels R. T-cell subset alterations in HIV-infected homosexual men: NIAID Multicenter AIDS cohort study. Clin Immunol Immunopathol. 1989;52(1):10–8. doi: 10.1016/0090-1229(89)90188-8

18. Liu C-M, Muirhead KA, George SP, Landay AL. Flow cytometric monitoring of human immunodeficiency virus-infected patients: simultaneous enumeration of five lymphocyte subsets. American journal of clinical pathology. 1989;92(6):721–8. doi: 10.1093/ajcp/92.6.721

19. Castro KG, Ward JW, Slutsker L, Buehler JW, Jaffe HW, Berkelman RL, et al. 1993 revised classification system for HIV infection and expanded surveillance case definition for AIDS among adolescents and adults. Clinical Infectious Diseases. 1993;17(4):802–10. doi: 10.1093/clinids/17.4.802

20. World Health Organization (WHO). Guidelines for managing advanced HIV disease and rapid initiation of antiretroviral therapy. Geneva, Switzerland: Licence: CC BY-NC-SA 3.0 IGO.; 2017 [Available from: https://www.who.int/publications/i/item/9789241550062]

21. Boulle A, Kenyon C, Skordis J, Wood R. Exploring the costs of a limited public sector antiretroviral treatment programme in South Africa. South African Medical Journal. 2002;92(10):811–7. doi:

22. Geffen N, Nattrass N, Raubenheimer C. The cost of HIV prevention and treatment interventions in South Africa. Faculty of Humanities, Centre for Social Science Research(CSSR): University of Cape Town (UCT),; 2003.

23. Stewart R, Loveday M. The operational plan: implementation of the antiretroviral therapy component: HIV and AIDS. South African health review. 2005;2005(1):224–46. doi:

24. South African Government. Operational plan for comprehensive HIV and AIDS care, management and treatment for South Africa 2003 [Available from: https://www.gov.za/sites/default/files/gcis_document/201409/aidsoperationalplan10.pdf]

25. Centers for Disease Control (CDC). Guidelines for the performance of CD4+ T-cell determinations in persons with human immunodeficiency virus infection 1992 [updated May 8. 41: RR-8:[1-17]. Available from: https://www.ncbi.nlm.nih.gov/pubmed/1350319]

26. Centers for Disease Control (CDC). Guidelines for the performance of CD4+ T-cell determinations in persons with human immunodeficiency virus infection. Recomm Rep 1992;41:1–17. 1992 [Available from: https://www.cdc.gov/mmwr/preview/mmwrhtml/00019952.htm]

27. Centers for Disease Control (CDC). 1993 revised classification system for HIV infection and expanded surveillance case definition for AIDS among adolescents and adults 1993 [Available from: https://www.ncbi.nlm.nih.gov/pubmed/1361652]

28. Centers for Disease Control (CDC). 1994 revised guidelines for the performance of CD4+ T-cell determinations in persons with human immunodeficiency virus (HIV) infections. Centers for Disease Control and Prevention 1994 [updated Mar 4. 43: RR-3:[1-21]. Available from: https://www.ncbi.nlm.nih.gov/pubmed/7908403]

29. Centers for Disease Control (CDC). 1997 revised guidelines for performing CD4+ T-cell determinations in persons infected with human immunodeficiency virus (HIV). Centers for Disease Control and Prevention 1997 [46: RR-2:[1-29]. Available from: https://www.cdc.gov/mmwr/preview/mmwrhtml/00045580.htm]

30. Republic of South Africa. National Health Laboratory Service Act 37 of 2000. Cape Town:; 2000.

31. Bateman C. Glencross confounds ‘wonder CD4’ critics. S Afr Med J. 2002;92(8):572–3. doi:

32. Barbesti S, Soldini L, Carcelain G, Guignet A, Colizzi V, Mantelli B, et al. A simplified flow cytometry method of CD4 and CD8 cell counting based on thermoresistant reagents: implications for large scale monitoring of HIV-infected patients in resource-limited settings. Cytometry B Clin Cytom. 2005;68(1):43–51. doi: 10.1002/cyto.b.20068

33. Barnett D, Walker B, Landay A, Denny TN. CD4 immunophenotyping in HIV infection. Nat Rev Microbiol. 2008;6(11 Suppl):S7–15. doi: 10.1038/nrmicro1998

34. Bergeron M, Nicholson J, Phaneuf S, Ding T, Soucy N, Badley A, et al. Selection of lymphocyte gating protocol has an impact on the level of reliability of T-cell subsets in aging specimens. Cytometry: The Journal of the International Society for Analytical Cytology. 2002;50(2):53–61. doi:

35. Mandy FF, Bergeron M, Minkus T. Evolution of leukocyte immunophenotyping as influenced by the HIV/AIDS pandemic: A short history of the development of gating strategies for CD4+ T-cell enumeration. Cytometry: The Journal of the International Society for Analytical Cytology. 1997;30(4):157–65. doi:

36. Glencross D, Scott LE, Jani IV, Barnett D, Janossy G. CD45-assisted PanLeucogating for accurate, cost-effective dual-platform CD4+ T-cell enumeration. Cytometry B Clin Cytom. 2002;50(2):69–77. doi: 10.1002/cyto.10068

37. Glencross DK, Stevens G, Scott LE, Mendelow BV, Stevens W. The challenge of laboratory monitoring of HIV. S Afr Med J. 2002;92(4):248. doi:

38. Glencross DK, Janossy G, Coetzee LM, Lawrie D, Scott LE, Sanne I, et al. CD8/CD38 activation yields important clinical information of effective antiretroviral therapy: findings from the first year of the CIPRA-SA cohort. Cytometry B Clin Cytom. 2008;74 Suppl 1:S131–40. doi: 10.1002/cyto.b.20391

39. Bateman C. Glencross confounds’ wonder CD4’critics. South African Medical Journal= Suid-afrikaanse Tydskrif vir Geneeskunde. 2002;92(8):572–3. doi:

40. Kestens L, Mandy F. Thirty-five years of CD4 T-cell counting in HIV infection: from flow cytometry in the lab to point-of-care testing in the field. Cytometry Part B: Clinical Cytometry. 2017;92(6):437–44. doi: 10.1002/cyto.b.21400

41. Coetzee LM, Glencross DK. Performance verification of the new fully automated Aquios flow cytometer PanLeucogate (PLG) platform for CD4-T-lymphocyte enumeration in South Africa. PLoS One. 2017;12(11):e0187456. doi: 10.1371/journal.pone.0187456

42. National Health Laboratory Service (NHLS). National CD4 Count Testing Programme 2022 [cited 2025 July]. Available from: https://www.nhls.ac.za/priority-programmes/cd4/]

43. Coetzee L-M, Glencross DK. Performance verification of the new fully automated Aquios flow cytometer PanLeucogate (PLG) platform for CD4-T-lymphocyte enumeration in South Africa. PLoS One. 2017;12(11):e0187456. doi: 10.1371/journal.pone.0187456

44. South African Government. Act no. 28 of 2024 - Public Procurement Act. Cape Town: GOVERNMENT GAZETTE, 23 July 2024; 2024.

45. Booplal N. Price List. In: Cassim N, editor.: National Health Laboratory Service; 2024.

46. Nedbank Group. Annual Exchange Rates: Nedbank Group; 2023 [cited 2024 25 February 2024]. Available from: https://www.nedbank.co.za/content/dam/nedbank/site-assets/AboutUs/Economics_Unit/Forecast_and_data/Daily_Rates/Annual_Average_Exchange_Rates.pdf]

47. National Health Laboratory Service (NHLS). Health Insurance (NHI) Bill Presentation: Parliamentary Portfolio CommitteeNational Health Laboratory Service (NHLS) Cape Town:; 2021 [Available from: https://nhi.healthjusticeinitiative.org.za/wp-content/uploads/2021/05/210520NHLS_Presentation-1.pdf?utm_source=copilot.com]

48. Parlimentary Monitoring Group (PMG). Medical Research Council; National Health Laboratory Services Annual Reports 2005/06 2007 [Available from: https://pmg.org.za/committee-meeting/8184/]

49. National Health Laboratory Service (NHLS). Annual Report 2019/20 Johannesburg, South Africa: National Health Laboratory Service (NHLS); 2020 [Available from: https://www.nhls.ac.za/wp-content/uploads/2021/03/NHLS_AR_2020_25_Nov.pdf]

50. Johnson LF, Dorrington RE. Modelling the impact of HIV in South Africa’s provinces: 2021 update. Centre for infectious Disease Epidemiology and Research working paper. 2023. doi:

51. Cassim N, Coetzee LM, Schnippel K, Glencross DK. Estimating implementation and operational costs of an integrated tiered CD4 service including laboratory and point of care testing in a remote health district in South Africa. PLoS One. 2014;9(12):e115420. doi: 10.1371/journal.pone.0115420

52. Glencross DK, Coetzee LM, Cassim N. An integrated tiered service delivery model (ITSDM) based on local CD4 testing demands can improve turn-around times and save costs whilst ensuring accessible and scalable CD4 services across a national programme. PLoS One. 2014;9(12):e114727. doi: 10.1371/journal.pone.0114727

53. Stevens WS, Cunningham B, Cassim N, Gous N, Scott LE. Cloud-based surveillance, connectivity, and distribution of the GeneXpert analyzers for diagnosis of tuberculosis (TB) and multiple-drug-resistant TB in South Africa. Molecular microbiology: diagnostic principles and practice. Third Edition 2016. p. 707–18.

54. Cohen L. The use and impacts of a Corporate Data Warehouse: The case of the national health laboratory service [Dissertation]: University of the Witwatersrand, Johannesburg (South Africa); 2018.

55. Glencross DK, Coetzee LM. Categorizing and Establishing CD4 Service Equivalency: Testing of Residual, Archived External Quality Assessment Scheme Sample Panels Enables Accelerated Virtual Peer Laboratory Review. Cytometry B Clin Cytom. 2019;96(5):404–16. doi: 10.1002/cyto.b.21772

56. Glencross DK, Janossy G, Coetzee LM, Lawrie D, Aggett HM, Scott LE, et al. Large-scale affordable PanLeucogated CD4+ testing with proactive internal and external quality assessment: in support of the South African national comprehensive care, treatment and management programme for HIV and AIDS. Cytometry Part B: Clinical Cytometry: The Journal of the International Society for Analytical Cytology. 2008;74(S1):S40–S51. doi: 10.1002/cyto.b.20384

57. Coetzee LM, Cassim N, Glencross DK. Newly implemented community CD4 service in Tshwaragano, Northern Cape province, South Africa, positively impacts result turn-around time. Afr J Lab Med. 2022;11(1):1376. doi: 10.4102/ajlm.v11i1.1376

58. Coetzee L, Cassim N, Glencross D. Implementation of a new’community’laboratory CD4 service in a rural health district in South Africa extends laboratory services and substantially improves local reporting turnaround time. South African Medical Journal. 2016;106(1):82–7. doi:

59. Carmona S, Bor J, Nattey C, Maughan-Brown B, Maskew M, Fox MP, et al. Persistent High Burden of Advanced HIV Disease Among Patients Seeking Care in South Africa’s National HIV Program: Data From a Nationwide Laboratory Cohort. Clin Infect Dis. 2018;66(suppl_2):S111–S7. doi: 10.1093/cid/ciy045

60. Coetzee LM, Cassim N, Glencross DK. Analysis of HIV disease burden by calculating the percentages of patients with CD4 counts <100 cells/microL across 52 districts reveals hot spots for intensified commitment to programmatic support. S Afr Med J. 2017;107(6):507–13. doi: 10.7196/SAMJ.2017.v107i6.11311

61. Coetzee LM, Cassim N, Sriruttan C, Mhlanga M, Govender NP, Glencross DK. Cryptococcal antigen positivity combined with the percentage of HIV-seropositive samples with CD4 counts <100 cells/μl identifies districts in South Africa with advanced burden of disease. PLoS One. 2018;13(6):e0198993. doi: 10.1371/journal.pone.0198993

62. Grimsrud A, Bygrave H, Doherty M, Ehrenkranz P, Ellman T, Ferris R, et al. Reimagining HIV service delivery: the role of differentiated care from prevention to suppression. Journal of the International AIDS Society. 2016;19(1):21484. doi:

63. National Department of Health (NDOH). National Antiretroviral Treatment Guidelines Pretoria, South Africa: National Department of Health (NDOH); 2004 [Available from: https://www.gov.za/sites/default/files/gcis_document/201409/artguidelines0.pdf]

64. National Department of Health (NDOH). Implementation of the universal test and treat strategy for HIV positive patients and differentiated care for stable patients Pretoria, South Africa: National Department of Health (NDOH); 2016 [Available from: https://sahivsoc.org/Files/22%208%2016%20Circular%20UTT%20%20%20Decongestion%20CCMT%20Directorate.pdf]

65. National Department of Health (NDOH). ART Clinical Guidelines for the Management of HIV in Adults, Pregnancy, Adolescents, Children, Infants and Neonates Pretoria, South Africa: National Department of Health (NDOH); 2019 [Available from: https://www.knowledgehub.org.za/system/files/elibdownloads/2020-05/2019%20ART%20Guideline%2028042020%20pdf.pdf]

66. National Department of Health (NDOH). ART Clinical Guidelines for the Management of HIV in Adults, Pregnancy and Breastfeeding, Adolescents, Children, Infants and Neonates Pretoria, South Africa: National Department of Health (NDOH); 2023 [Available from: https://knowledgehub.health.gov.za/system/files/elibdownloads/2023-07/National%20ART%20Clinical%20Guideline%20AR%204.5%2020230713%20Version%204%20WEB.pdf]

67. Calmy A, Ford N, Hirschel B, Reynolds SJ, Lynen L, Goemaere E, et al. HIV viral load monitoring in resource-limited regions: optional or necessary? Clinical infectious diseases. 2007;44(1):128–34. doi:

68. Peter T, Ellenberger D, Kim AA, Boeras D, Messele T, Roberts T, et al. Early antiretroviral therapy initiation: access and equity of viral load testing for HIV treatment monitoring. The Lancet Infectious Diseases. 2017;17(1):e26–e9. doi:

69. Scott LE, Glencross DK. Monitoring reproducibility of single analysis, single platform CD4 cell counts in a high volume, low resource laboratory setting using sequential bead count rates. Cytometry B Clin Cytom. 2005;67(1):31–2. doi: 10.1002/cyto.b.20066

70. Lawrie D, Coetzee LM, Glencross DK. A model for continuous quality control incorporating sample-to-sample assessment of optical alignment, fluorescence sensitivity, and volumetric operation of flow cytometers. Cytometry B Clin Cytom. 2010;78(3):201–10. doi: 10.1002/cyto.b.20520

71. Cassim N, Coetzee LM, Tepper MEE, Perelson L, Glencross DK. Timely delivery of laboratory efficiency information, Part II: Assessing the impact of a turn-around time dashboard at a high-volume laboratory. Afr J Lab Med. 2020;9(2):948. doi: 10.4102/ajlm.v9i2.948

72. Cassim N, Tepper ME, Coetzee LM, Glencross DK. Timely delivery of laboratory efficiency information, Part I: Developing an interactive turnaround time dashboard at a high-volume laboratory. Afr J Lab Med. 2020;9(2):947. doi: 10.4102/ajlm.v9i2.947

73. Coetzee LM, Cassim N, Glencross DK. Using laboratory data to categorise CD4 laboratory turn-around-time performance across a national programme. Afr J Lab Med. 2018;7(1):665. doi: 10.4102/ajlm.v7i1.665

74. Cassim N, Coetzee LM, Stevens WS, Glencross DK. Addressing antiretroviral therapy-related diagnostic coverage gaps across South Africa using a programmatic approach. Afr J Lab Med. 2018;7(1):681. doi: 10.4102/ajlm.v7i1.681

75. Larson BA, Rockers PC, Bonawitz R, Sriruttan C, Glencross DK, Cassim N, et al. Screening HIV-Infected Patients with Low CD4 Counts for Cryptococcal Antigenemia prior to Initiation of Antiretroviral Therapy: Cost Effectiveness of Alternative Screening Strategies in South Africa. PLoS One. 2016;11(7):e0158986. doi: 10.1371/journal.pone.0158986

76. Cassim N, Coetzee LM, Schnippel K, Glencross DK. Estimating the cost-per-result of a national reflexed Cryptococcal antigenaemia screening program: Forecasting the impact of potential HIV guideline changes and treatment goals. PLoS One. 2017;12(8):e0182154. doi: 10.1371/journal.pone.0182154

77. Cassim N, Coetzee LM, Govender NP, Glencross DK. District and sub-district analysis of cryptococcal antigenaemia prevalence and specimen positivity in KwaZulu-Natal, South Africa. Afr J Lab Med. 2018;7(1):757. doi: 10.4102/ajlm.v7i1.757

78. Govender NP, Glencross DK. National coverage of reflex cryptococcal antigen screening: A milestone achievement in the care of persons with advanced HIV disease. S Afr Med J. 2018;108(7):534–5. doi: 10.7196/SAMJ.2018.v108i7.13094

79. Bergeron M, Daneau G, Ding T, Sitoe NE, Westerman LE, Stokx J, et al. Performance of the PointCare NOW system for CD4 counting in HIV patients based on five independent evaluations. PLoS One. 2012;7(8):e41166. doi: 10.1371/journal.pone.0041166

80. Van Turha L, Maharaj K, Rose A, Boeke C, Peter TF, Vojnov L, et al. Point-of-care CD4+ technology implementation in Free State, South Africa, was associated with improved patient health outcomes. S Afr Med J. 2020;110(2):126–31. doi: 10.7196/SAMJ.2020.v110i2.13823

81. Glencross DK, Coetzee LM, Faal M, Masango M, Stevens WS, Venter WF, et al. Performance evaluation of the Pima point-of-care CD4 analyser using capillary blood sampling in field tests in South Africa. J Int AIDS Soc. 2012;15(1):3. doi: 10.1186/1758-2652-15-3

82. Faal M, Naidoo N, Glencross DK, Venter WD, Osih R. Providing immediate CD4 count results at HIV testing improves ART initiation. J Acquir Immune Defic Syndr. 2011;58(3):e54–9. doi: 10.1097/QAI.0b013e3182303921

83. Coetzee LM, Moodley K, Glencross DK. Performance Evaluation of the Becton Dickinson FACSPresto Near-Patient CD4 Instrument in a Laboratory and Typical Field Clinic Setting in South Africa. PLoS One. 2016;11(5):e0156266. doi: 10.1371/journal.pone.0156266

84. Sloot R, Glenshaw MT, van Niekerk M, Meehan SA. Rapid point-of-care CD4 testing at mobile units and linkage to HIV care: an evaluation of community-based mobile HIV testing services in South Africa. BMC Public Health. 2020;20(1):528. doi: 10.1186/s12889-020-08643-3

85. Vojnov L, Markby J, Boeke C, Harris L, Ford N, Peter T. POC CD4 testing improves linkage to HIV care and timeliness of ART initiation in a public health approach: a systematic review and meta-analysis. PLoS One. 2016;11(5):e0155256. doi:

86. Rosen S, Ketlhapile M, Sanne I, DeSilva MB. Cost to patients of obtaining treatment for HIV/AIDS inSouth Africa. South African Medical Journal. 2007;97(7):524–9. doi:

87. South African Government. Act no.1 of 1999 - Public Finance Management Act (PFMA). Cape Town: GOVERNMENT GAZETTE, 1999; 1999.

88. Joint United Nations Programme on HIV/AIDS (UNAIDS). Impact of US funding freeze on HIV response in South Africa 2025 [Available from: https://www.unaids.org/en/resources/presscentre/featurestories/2025/march/20250307_South-Africa_fs]

89. Nxumalo CT, Buthelezi U, Chiya H, Makgobole MU, Mpofana N, Mgwaba T, et al. Achieving the fast track 90-90-90 and 95-95-95 targets in sub-Saharan Africa: A rapid review. J Public Health Afr. 2025;16(1):691. doi: 10.4102/jphia.v16i1.691

